# A standardised test to evaluate audio-visual speech intelligibility in French

**DOI:** 10.1101/2023.01.18.23284110

**Authors:** Loïc Le Rhun, Gerard Llorach, Tanguy Delmas, Clara Suied, Luc H Arnal, Diane S Lazard

## Abstract

**Objective:** Lipreading, which plays a major role in the communication of the hearing impaired, lacked a French standardised tool. Our aim was to create and validate an audio-visual (AV) version of the French Matrix Sentence Test (FrMST).

**Design:** Video recordings were created by dubbing the existing audio files.

**Sample:** Thirty-five young, normal-hearing participants were tested in auditory and visual modalities alone (Ao, Vo) and in AV conditions, in quiet, noise, and open and closed-set response formats.

**Results:** Lipreading ability (Vo) varied from 1% to 77%-word comprehension. The absolute AV benefit was 9.25ℒdB SPL in quiet and 4.6ℒdB SNR in noise. The response format did not influence the results in the AV noise condition, except during the training phase. Lipreading ability and AV benefit were significantly correlated.

**Conclusions:** The French video material achieved similar AV benefits as those described in the literature for AV MST in other languages. For clinical purposes, we suggest targeting SRT80 to avoid ceiling effects, and performing two training lists in the AV condition in noise, followed by one AV list in noise, one Ao list in noise and one Vo list, in a randomised order, in open or close set-format.

## Introduction

During the COVID pandemic, the general population became aware of the importance of lipreading even among normal-hearing listeners, when wearing facial masks ^[4]^. Several studies have demonstrated that the combination of masks and background noise has a negative impact on speech intelligibility, even in normal listeners ^[5–7]^. Yi et al.^[7]^ showed a decrease of 20% of sentence comprehension in normal listeners when adding a mask to the locutor. These results underpin the limits of understanding speech in adverse listening conditions when relying on the auditory modality alone (unimodal condition, Ao). Thus, visual cues from lips’ movements and facial expressions help disambiguate the auditory input ^[8–11]^. Visemes are known to play a major role in the communication of the hearing impaired ^[2]^ and to have a predictive value in hearing rehabilitation by cochlear implants ^[3]^ or conventional hearing aids_[1]_.

While many tests assessing speech understanding in Ao are available and validated in the international literature, calibrated automatized audio-visual (AV) testing material is scarce ^[12,13]^ and does not exist in French. Adding an AV assessment tool thus seems important to explore deafness and its compensation, and more generally AV synergy in deprived and non-deprived participants.

A wide variety of speech audiometry tests are available in French, from words in quiet (e.g., Fournier^[14]^, Lafon^[15]^) to sentences in noise (e.g., MRT: Modified Rhyme Test ^[16]^, FrMST: French Matrix Sentence Test ^[17]^, VRB: Vocale Rapide dans le Bruit ^[18]^). The FrMST, which is the French adaptation of the international MST, caught our attention because it has been widely used in clinical practice in France since its validation in 2012 ^[17]^, and is available in different languages (for comparison purposes). Its stereotypical pattern –28 lists of 10 sentences of 5 words, each generated from 50 words (10 nouns + 10 verbs + 10 numbers + 10 objects + 10 colours)– is of interest when performing neurofunctional imaging, such as EEG ^[19]^. In practice, the MST uses a staircase procedure presenting the sentences in a fixed background noise ^[20,21]^. The signal presentation level varies to provide a specified speech reception threshold (SRT) (e.g., 50%).

Four teams have already validated an audio-visual version of the MST into their respective languages (New Zealander English, Malay, Dutch, and German) ^[22–24,13,12]^. Instead of recreating the audio-visual material, Llorach et al. ^[12]^ dubbed the original German MST audio sentences. Recording new MST speech material from scratch is a demanding process, which can induce differences in speech perception of up to 6 dB SNR compared to previous versions, depending on the talker ^[25]^. Therefore, dubbing the original validated audio sentences with videos ensures results more comparable to those of the literature. To avoid AV lags, Llorach et al.^[12]^ described the protocol for recording and selecting the best aligned videos from their audio recordings ^[12]^ (see the paragraph "Selection of the videos” for further details).

Here, we reproduced and improved the methodology (creation and validation) of Llorach et al. ^[12]^, and compared our results in terms of AV training effect, test-retest variability, lipreading ability in the visual only (Vo) condition, and AV gain in young normal hearing participants.

## Material and Method

All the device references used in this study are given in Table 1.

**Table 1:**
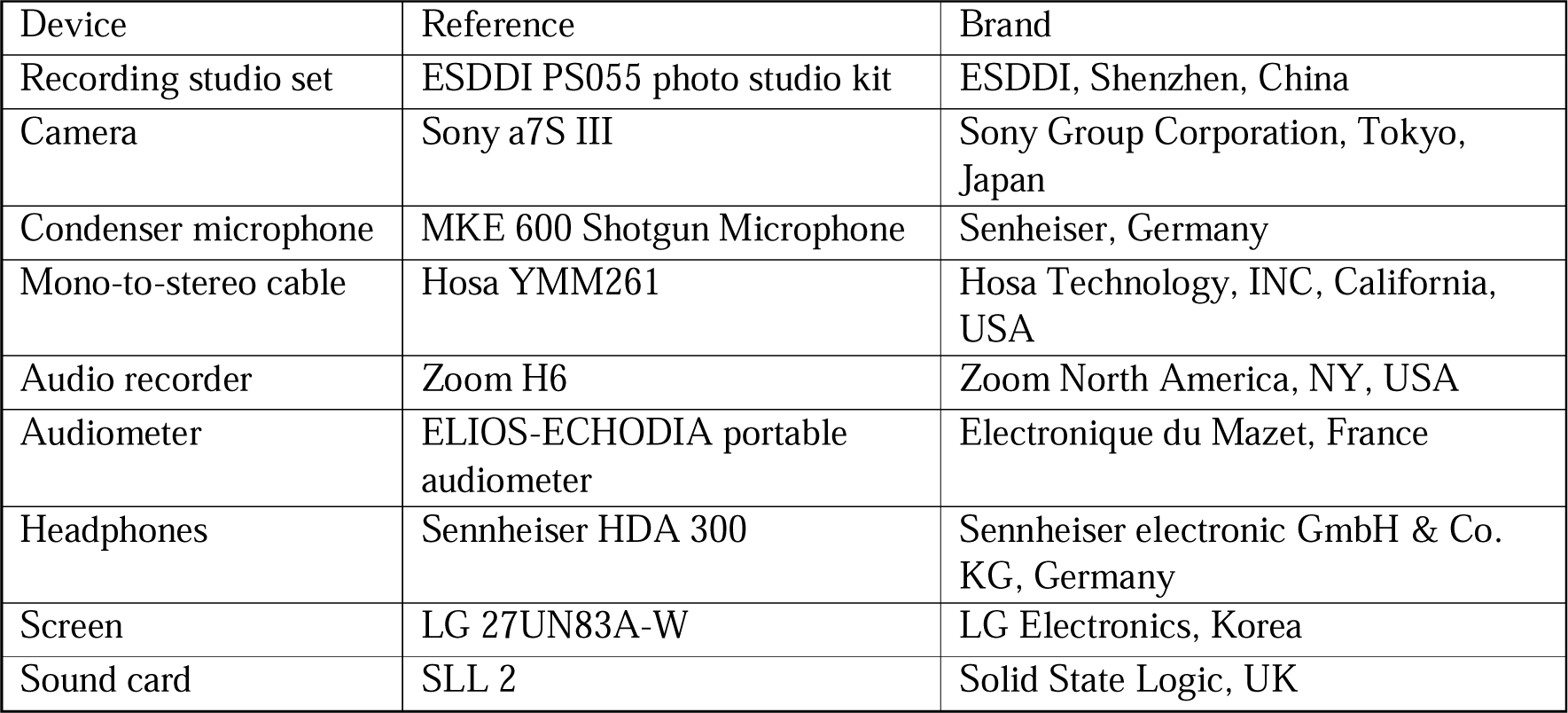
References of the devices used.

Scripts and guidelines for automatically recording, processing and cutting video material are available in the GitHub repository, at the following address: https://github.com/gerardllorach/audiovisualdubbedMST/.

### Dubbing the auditory material and recording the videos

The French version was validated in 2012 ^[17]^ and is largely used in clinical practice. The initial corpus included 280 sentences. We manually discarded the sentences for which the original audio recording presented artifacts due to the merging of different sentences. From the remaining 150 sentences, forty-five lists of 20 sentences were created. In each list of 20 sentences, no word appeared more than twice and no sentence was repeated. The same female speaker who originally recorded the auditory French version participated in the video recordings.

The videos were recorded in an anechoic room, transformed into a film studio (Figure 1). The set up followed the same principle as in Llorach et al. (2022). The speaker was filmed while repeating the sentence she was hearing through an earphone placed in her left ear. The camera recorded the video together with the two audio tracks (original sentence and microphone signal) separately. The camera recorded images at 50 fps/full HD and speech with a 48 kHz sampling rate and a 16-bit LPCM (linear pulse-code modulation) sampling format. Each sentence was presented 4 times in a row to the speaker. The speaker first heard three pure tones through the earphone to prepare herself and listened to the first presentation to know the five words to repeat. She then had to dub the three following sentences while simultaneously hearing them. The requirements were a neutral face, without exaggeration of the articulation, while keeping her eyes fixed and focused on the camera and avoiding blinking. Several takes could be recorded for the same sentence.

**Figure 1:**
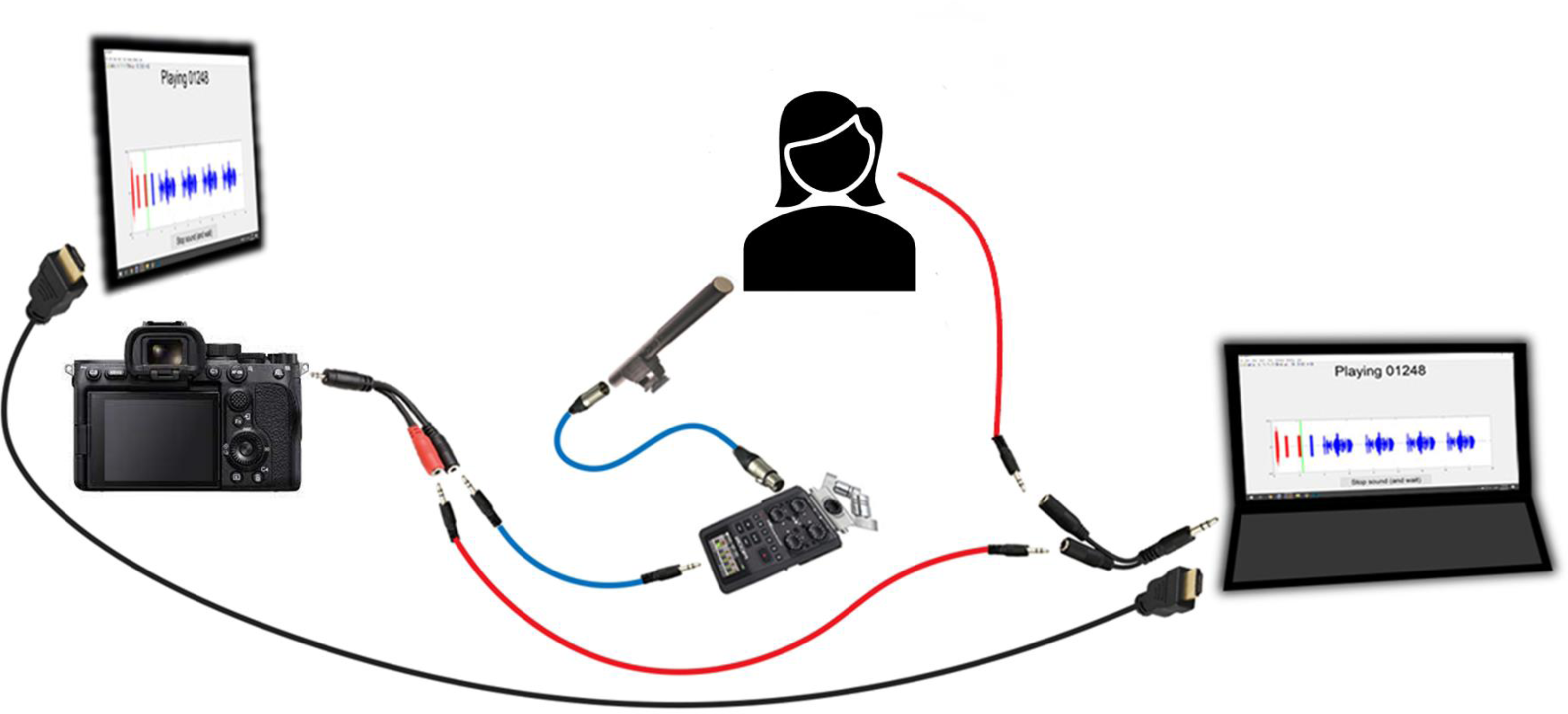
Schematic drawing of the recording setup: the camera and the screen showing the visual aid are on the left side. The audio input of the camera is connected to the mono-to-stereo jack adapter. The input of this adapter is the recorded signal (blue path) and the original speech (red path). The speaker with a green background and an earphone playing the original speech signal is in the center. The condenser microphone, below the speaker, is connected to the audio recorder. The output of the audio recorder goes to the mono-to-stereo adapter. On the right: the computer that generates the three tones and the sentence repetitions (waveforms). The audio output of the computer is connected to the earphone and to the mono input of the mono-to-stereo adapter. An HDMI cable connects the computer screen to the visual aid on top of the camera to duplicate the screen.

The resulting video files, which contained the visual speech, the original audio sentences of the French MST and the recorded speech were processed automatically via open-source scripts. During the recording session, the camera recorded continuously i.e., a video file may contain several takes of sentences. The duration of the sentence was retrieved from the original MST audio files and each take was cut into repetitions (three sentence repetitions for each take).

The MATLAB scripts used and guidelines to record the video material can be found in the GitHub repository (see Statements).

### Selection of the videos

Videos in which the expressions were not neutral were manually removed. To select the best videos, i.e. those most synchronized with the original audio, we quantified the time misalignments between the original auditory sentences and the sentences dubbed by the speaker during the recording, using the same approach as in Llorach et al.^[12]^, which relies on the dynamic time warping (DTW) algorithm ^[27]^. In technical terms, the DTW provides a warping path that can be interpreted as a time misalignment showing words spoken too early or too late and/or words spoken slower or faster when comparing two sentences. The time misalignment score allows selecting the best match between the recordings and the original sentences, enabling the selection of the most synchronized videos with the original audio (the new audio recordings are only used to select the most suitable videos). We computed the mel-spectrograms of the original and the dubbed sentences and applied the DTW algorithm. We obtained the selection of the best aligned sentences. We secondarily constrained the algorithm to compare the mel spectrogram of one speech signal of the recorded sentences to a maximum of three frames of the matching original audio recording, in order to provide a more accurate time misalignment score. Figure 2 reports the time misalignment score of one sentence, i.e. the average temporal difference between the reference and the best recorded sentence (go to https://doi.org/10.5281/zenodo.8188917 for all sentences). Figure 3 shows the distribution of the time misalignment scores of the original sentences against their final best selected sentences (left box plot: median 32.2 ms, mean 32.8 ± 9.5 ms, range: 16.4 to 59.3), against their four repetitions (middle box plot: mean 38.4 ± 14.6 ms) and against mismatched sentences (right box plot: mean 114.3 ± 96 ms). The ideal, but unrealistic, value would be a time misalignment score of 0 ms. The extreme case (no alignment) is displayed by the third box plot iteratively fitting mismatched sentences. The significant improvement in score between the three calculations (p<10^-3^) shows the relevance of the DTW method.

**Figure 2:**
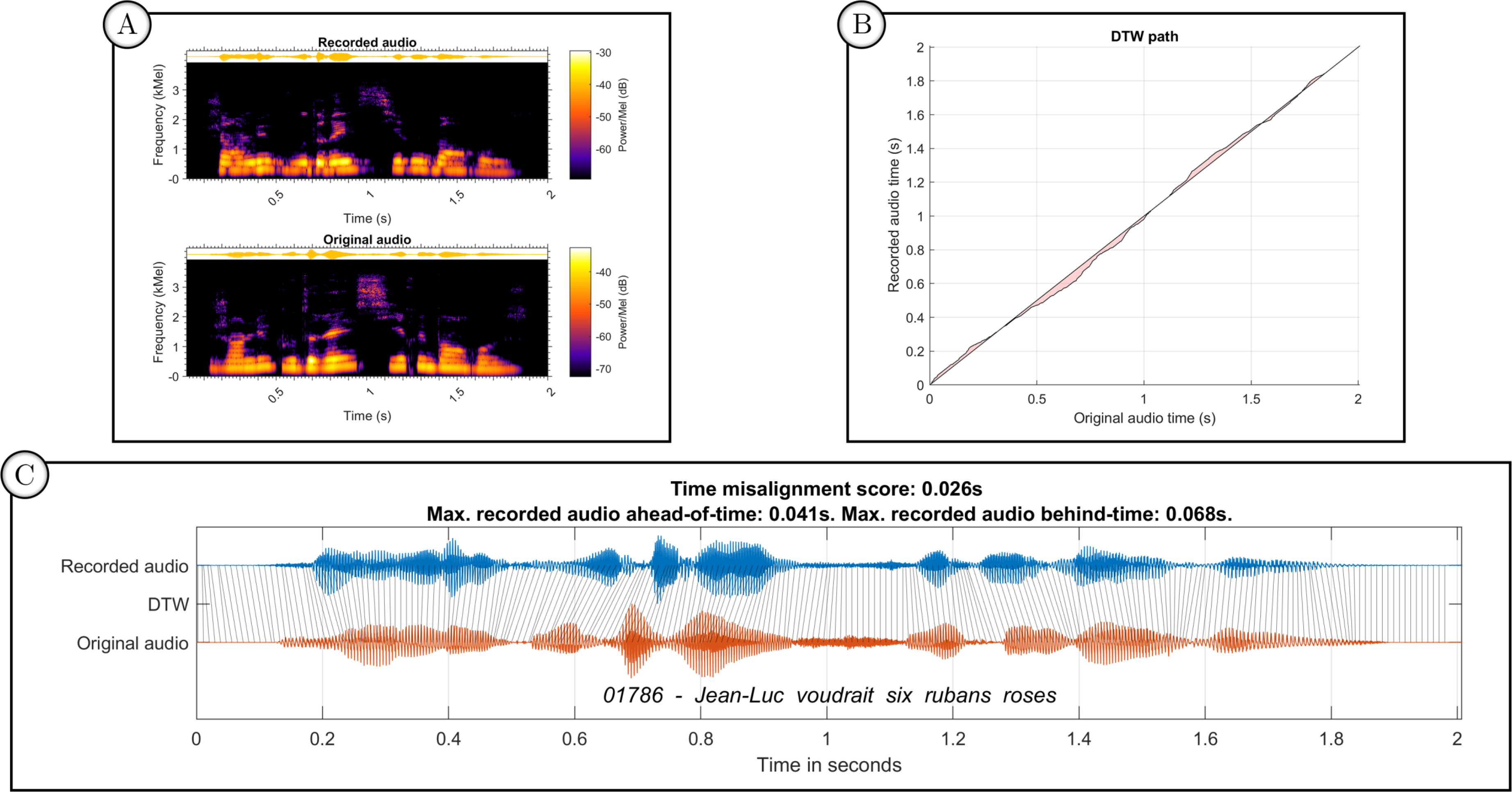
Dynamic time wrapping (DTW) between an original audio and its most synchronous recording. Panel A shows the mel spectrograms of the two signals, which are used to compute the DTW. Panel B shows the DTW path between the mel spectrograms displayed in Panel A. The red area represents the time misalignment between the two spectrograms. It is used to compute the time misalignment score and maximum time misalignments (recorded audio ahead-of-time and behind-of-time) displayed in Panel C (top). Panel C shows the waveforms of the recorded audio (blue), the original audio (orange) and their temporal misalignment according to the frame-restricted DTW. The black lines in between the two signals, labelled as DTW, show the match between mel spectrogram frames. The sentence code and spoken words are shown below the signals (they are not temporally aligned with the spectrograms).

**Figure 3:**
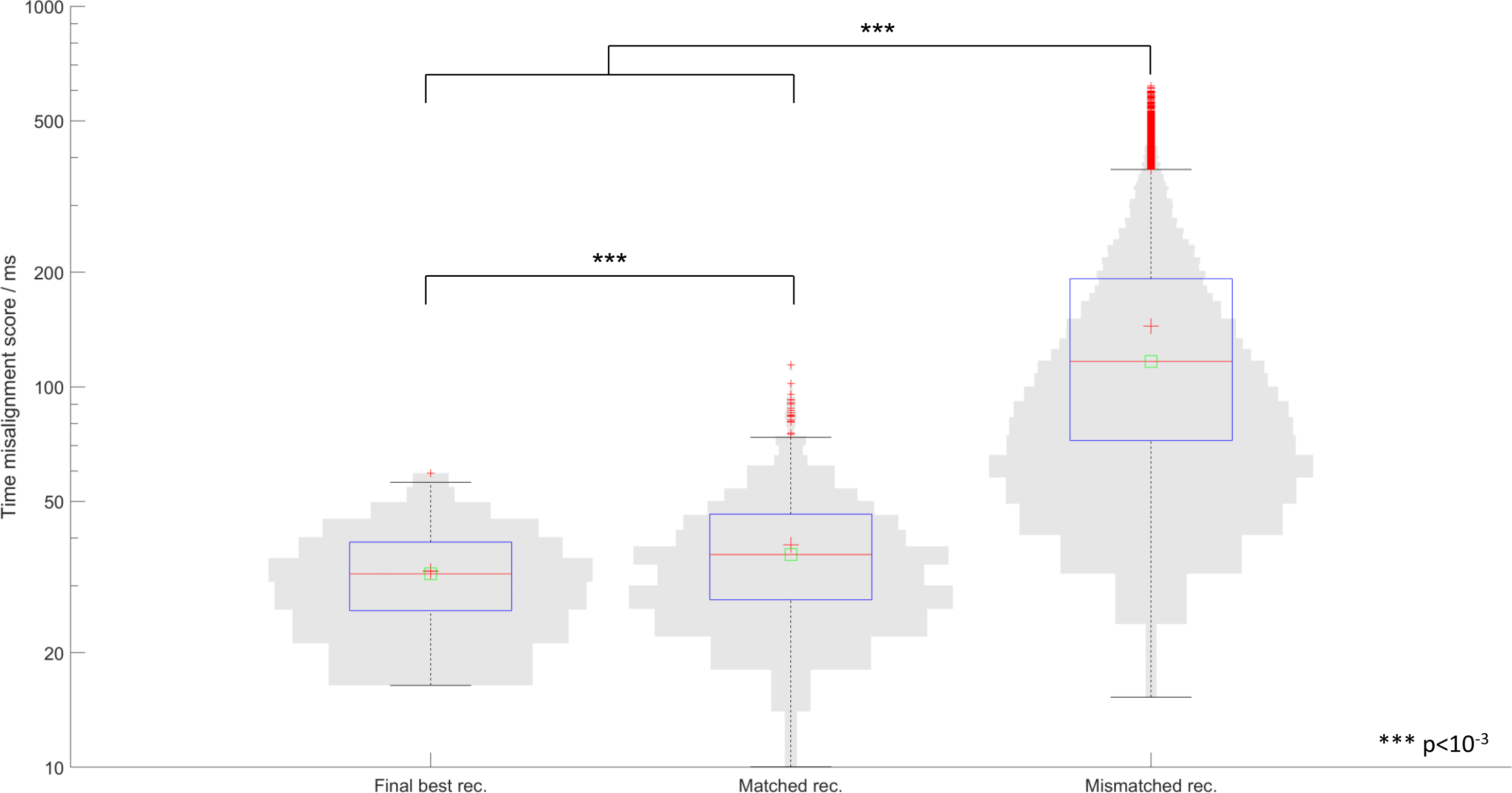
Relevance and performance of the DTW method for selecting the best recorded audio tracks The left box plot displays the time misalignment scores between the original sentences and their best-matched recordings (n=150 misalignment scores), the middle box plot displays the time misalignment scores between the original sentences and their 4 recordings (n=150 x 4 misalignment scores), the right box plot displays the time misalignment scores between the original sentences and the mismatched final best recordings (n=150□×□149 misalignment scores; the comparison between the sentence and its matching recording has been removed). The vertical axis is on a logarithmic scale. The large red cross represents the mean, the small red crosses represent outliers, and the red line represents the median. The lower whisker represents the 1^st^ percentile, and the upper whisker represents the 99^th^ percentile. The lower the score, the more aligned the audio signals.

To summarize, the final audio-visual material consisted of the original validated auditory sentences of the FrMatrix and their best synchronized videos (selected on the smallest time misalignment scores between the two audio tracks). No participant reported any audio-visual lag during the test.

### Validation of the AV material

#### Participants

This project was approved by the Ethics committee CPP Tours-Region Centre-Ouest 1 (project identification number 2020T317 RIPH3 HPS). All the participants gave their written consent and received financial compensation.

The test was validated on a gender-balanced cohort of 35 normal-hearing participants. Two participants did not perform the retest session. Participants were 20 to 29 years old (mean age: 24.4 years with a standard deviation (SD) of 2.8), with no known hearing problems or difficulty understanding in noisy environments. They had normal or corrected vision and no specific lipreading skills. All participants were native French speakers and their level of education ranged from 0 to 8 years after high school (mean: 4 years). Their assessed mean pure tone average (PTA) was 1.22 dB HL (range: −4.25; 5).

#### Set up

The experiment was performed in an anechoic booth. Participants watched videos on a monitor at a viewing distance of 80 cm and listened to the speech sounds through binaural audiometric headphones. The recorded videos and the original auditory sentences were played using VLC® (version 3.0.16). The acoustic signal was sent with a SLL 2 sound card. The acoustic levels were calibrated using an artificial ear connected to a sound level meter.

#### Stimuli

Each video had a green background and started with a 500 ms still face. The participants were tested in quiet and in noise with the long-term stationary noise of the average speech spectrum (LTASS) from the original French MST ^[17]^, according to a usual adaptive procedure ^[20,21]^

In quiet, the presentation level of the sentences started at 25 dB SPL, except for the first tested participant. In noise, the presentation level of the sentences started at 60 dB SPL, with a fixed noise of 65 dB SPL.

#### Experimental conditions

Three different modalities were tested, Audio only (Ao), Visual Only (Vo) and Audio-visual (AV). Ao and AV modalities were tested in quiet and in noise, in open-set and closed-set response formats (8 conditions). The Vo modality was presented in noise to be more ecological and in closed-set response format (open-set format being too difficult for naive participants). In the open-set format, participants repeated aloud what they understood. In the closed-set format, participants selected their responses on a matrix between all the 50 possible words and a "no response" option.

For each condition, the participant was tested with one list of 20 sentences. The adaptive procedure was set to reach an individual Speech Reception Threshold of 80% (SRT80) to avoid ceiling effects in the AV conditions. Indeed, some participants are able to understand more than 50% of the content only relying on lipreading ^[13]^.

SRT80 is expressed in dB SPL in quiet conditions and in dB SNR (signal-to-noise ratio) in noisy conditions. For the Vo condition, we report the percentage of words correctly understood. No feedback was provided.

#### Procedure

A test-retest procedure (two sessions) was performed, spaced between 3 to 59 days apart.

All participants undertook a training phase before each session to control for the learning effect. This training phase was composed of four AV lists in noise during the first session, and one AV list during the retest session (Figure 4). The response format (open-set or closed-set) of the training lists was randomly assigned to each participant. This response format was the same for both training phases. Participants started the session with the response format assigned during the training, and secondarily performed the same conditions with the other response format.

**Figure 4:**
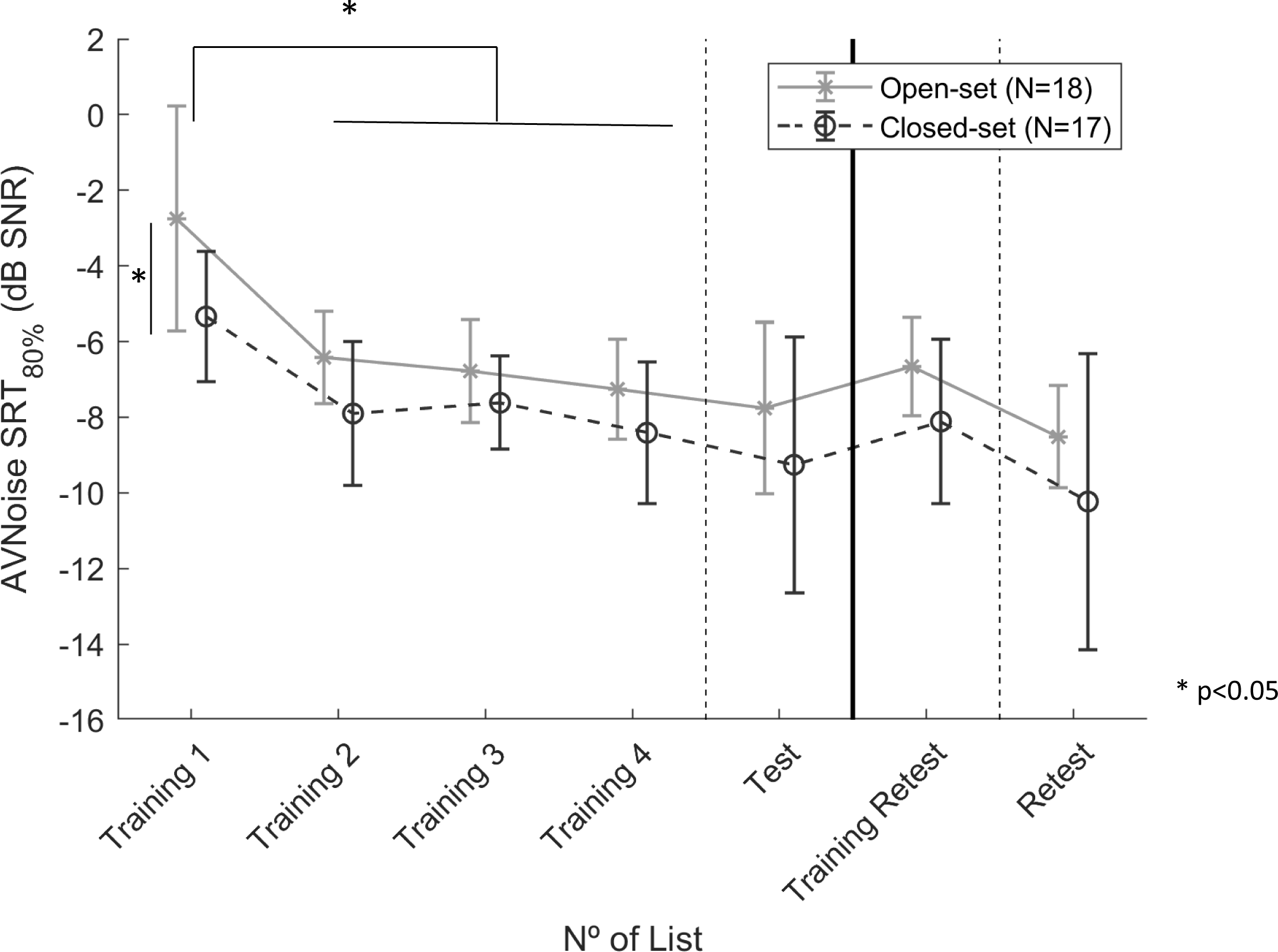
Audio-visual training effect in noise during the test and retest sessions Four training lists were used during the test session. One list was used during the retest session. Results are expressed in mean SRT ± SD, with black dashed lines and circles for the participants who performed the training in closed-set response format, and continuous grey line for those who performed the training in open-set response format. *: p <10^-3^. According to Bonferroni *post hoc* tests; SRTs of Training 1 were significantly different from the SRTs of the three other training lists regardless of response formats. SRTs of Training 1 were significantly different between response formats

The order of the modalities (AV, Ao, Vo) and the environment (quiet and noise) was randomised for the two sessions. All participants performed the conditions in quiet and in noise.

We added another condition to the second session to compare our results with those described in Jansen et al. ^[17]^, i.e. SRT50 in Ao in noise and closed-set format. This measure was performed after the training list.

#### Statistical analysis

Results are expressed in means ± standard deviation (SD). ANOVAs for repeated measures were performed using JASP (v0.16.3, University of Amsterdam, Netherlands), followed by *post-hoc* multiple comparisons with Bonferroni corrections. If the sphericity assumption was not met, a Greenhouse Geisser correction was used. We searched for correlations (Pearson) between the Vo scores (lipreading skills) and the AV benefit (AV – Ao scores).

## Results

### SRT50

In 2012, Jansen et al. ^[17]^ validated the French version of the MST with a mean SRT50 of −6 dB SNR (±0.6). Here, we obtained a mean SRT50 of −7.75 dB SNR (±0.79).

### Evaluation of the training effect

The clinical use of the MST starts with a systematic training session based on two training lists to get used to the type of material and background noise ^[17]^. For the AV version, we tested four lists during the training phase of the test session and one list during the retest session (AV Noise condition, closed or open-set format). The test and retest sessions were spaced 17 days apart on average (range: 3-59).

Figure 4 shows the training effect (mean SRT80% in dB SNR for the AV condition in noise in the response format performed) during the test and retest sessions. On average, the training effect between the first training list and the test was −4.5 dB SNR for the first session, and - 1.93 dB SNR for the second session, regardless of the response format. The repeated-measures ANOVA with a Greenhouse-Geisser correction (Maulchy’s test X^2(5) = 22.4, p <10^-3^) showed a significant effect of training lists (F(2, 65.90) = 51.7, p <10^-3^) independently of the response format. *Post-hoc* multiple comparisons with Bonferroni corrections showed that this effect came from the difference between the first list of the first session and the three other training lists (p<10^-3^, Figure 4 subsequent SRTs of the remaining training lists did not significantly differ. The analysis further showed a significant improvement during the second session between the training and the test (F(1, 31) = 25, p <10^-3^). There was a significant score difference of 1.5 dB SNR (±0.4) between the open-set and closed-set formats during the first training session only (F(1, 33) = 11.6, p = 0.002). The interaction with the number of training lists was not significant (F(2, 65.90) = 2.58, p = 0.084)

### Audio-only (Ao) and Audio-visual evaluation

Table 2 shows the mean SRTs (±SD) obtained in the different Ao and AV conditions (noise/quiet, closed/open response format) averaging test and retest results (except for the two participants who only performed the first session). There was no effect of the response formats.

**Table 2:** Mean Speech Reception Thresholds SRT80% (±Standard Deviation) obtained in the different Audio-only (Ao) and Audio-visual (AV) conditions, averaging test and retest results.

| Mean SRT (dB SNR) |  | Mean SRT (dB SPL) |  |
| --- | --- | --- | --- |
| Ao Noise Closed | -4,8 (0,9) | Ao Quiet Closed | 25,4 (3,2) |
| Ao Noise Open | -4,6 (1,1) | Ao Quiet Open | 26,6 (3,8) |
| AV Noise Closed | -9,5 (3,2) | AV Quiet Closed | 16,4 (5,6) |
| AV Noise Open | -8,9 (2,5) | AV Quiet Open | 17,1 (5,1) |

SRT80% expressed in dB SNR for the conditions in noise and in dB SPL for the conditions in quiet. For the conditions in quiet, we use the term SRT80% by extension, but the percentage expresses the sound presentation level for 80% word recognition.

The absolute SRT benefit obtained by adding the visual modality to the auditory alone condition (AV - Ao) was 4.6 dB SNR in noise and 9.25 dB SPL in quiet.

The AV conditions allowed some participants to reach the SRT80 with speech presentation levels below 0 dB SPL in quiet and SNRs below −20 dB SNR in noise (Figure 5). These auditory presentation levels are near the threshold of human hearing and speech detection ^[26]^. Thus the speech detection threshold (SDT) of the female German MST has been evaluated at - 16.9 dB SNR in the Ao condition in noise ^[27]^, a threshold that can be theoretically lowered by −3 dB when adding visual cues, as demonstrated by Bernstein et al ^[28]^. We believe that these participants reached these presentation levels because they were able to lipread more than 80% of the material, forcing the adaptive procedure to lower the SNRs or SPLs. As Llorach et al. ^[12]^ did, we considered that these presentations levels were not fully representative of audio-visual speech reception and limited their SRT scores to −20 dB SNR and 0 dB SPL. This score limitation concerned 3 scores from 3 participants out of 445 SRTs (0.7% *versus* 5% in Llorach et al. ^[12]^). It is to note that Ross et al ^[29]^ and Sumby and Pollack ^[30]^ found audio-visual speech reception thresholds below –20 dB SNR. However, the material used was different (noise and target sentences) making the comparison more difficult than with the German MST version that used similar testing conditions. Future work should consider measuring the real SDT of the French MST.

**Figure 5:**
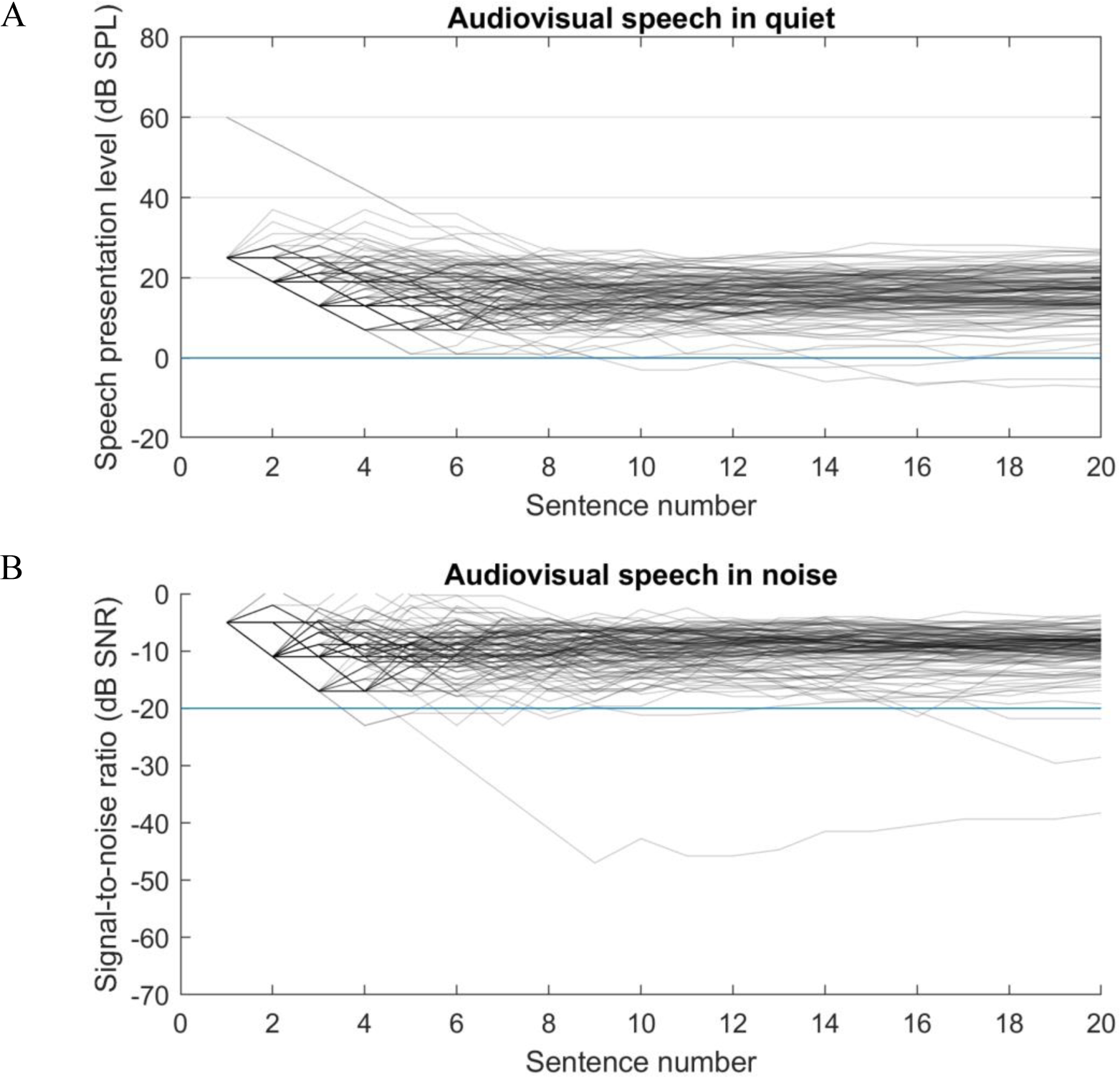
Adaptive speech presentation levels for the Audio-visual conditions in quiet (a) and adaptive signal to noise ratios for the Audio-visual conditions in noise (b) The adaptive procedure changed the speech levels to reach 80% of intelligibility (SRT80 for a fixed background noise). Each line shows a single list of 20 sentences per participant. Below the horizontal line at 0 dB SPL (a) and at −20 dB SNR (b), participants understood speech using only visual cues. The presentation level in quiet started at 25 dB SPL, except for the first participant of the study who started with a presentation level of 60 dB.

### Lipreading (Vo) and audio-visual benefit

Participants had a wide range of lipreading abilities (Figure 6). The individual Vo scores in noise and closed-set format ranged from 1% to 77%-word intelligibility, with an average of 51.3% (±16.2%), in accordance with that of Llorach et al. ^[12]^ in the same testing conditions (50% ±21.4%). There was an average intelligibility improvement between the test and the retest sessions of 4.8 % (i.e., 5 words over 100 presented during the 20 sentences). This improvement was significant (paired t-test: p<10^-3^). Lipreading scores (Vo) correlated with the AV benefit (p <10^-3^) in all AV conditions (Figure 7). The Pearson r ranged from −0.55 (in quiet and open-set format) to −0.74 (in noise and open-set format).

**Figure 6:**
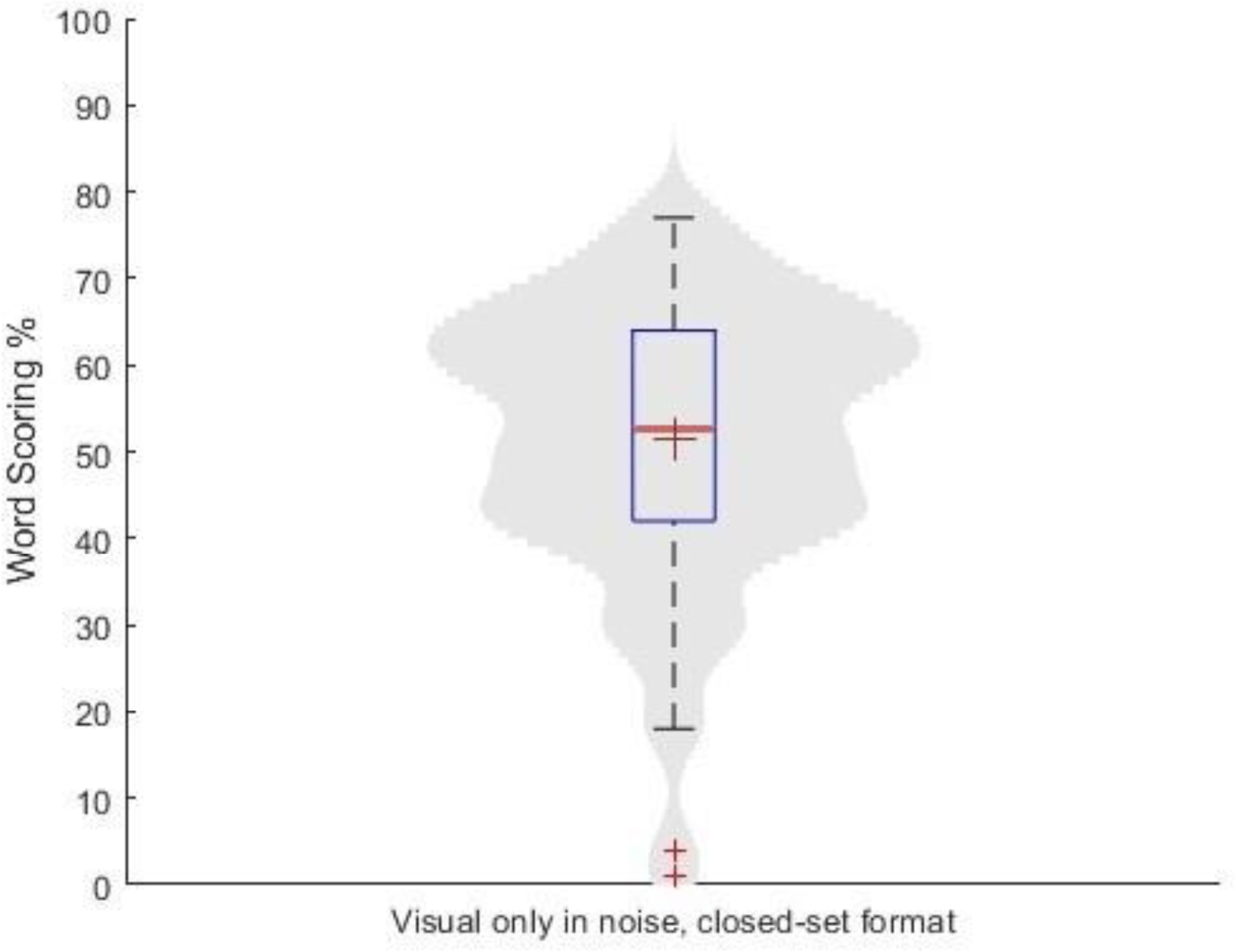
Boxplot and distribution of the lipreading scores: Visual only in noise and closed-set format The mean and the median are represented by a red cross and a red line, respectively. The outliers are represented by smaller red crosses. The boxplot is smoothed with the Matlab Kdensity function that returns a probability density estimate.

**Figure 7:**
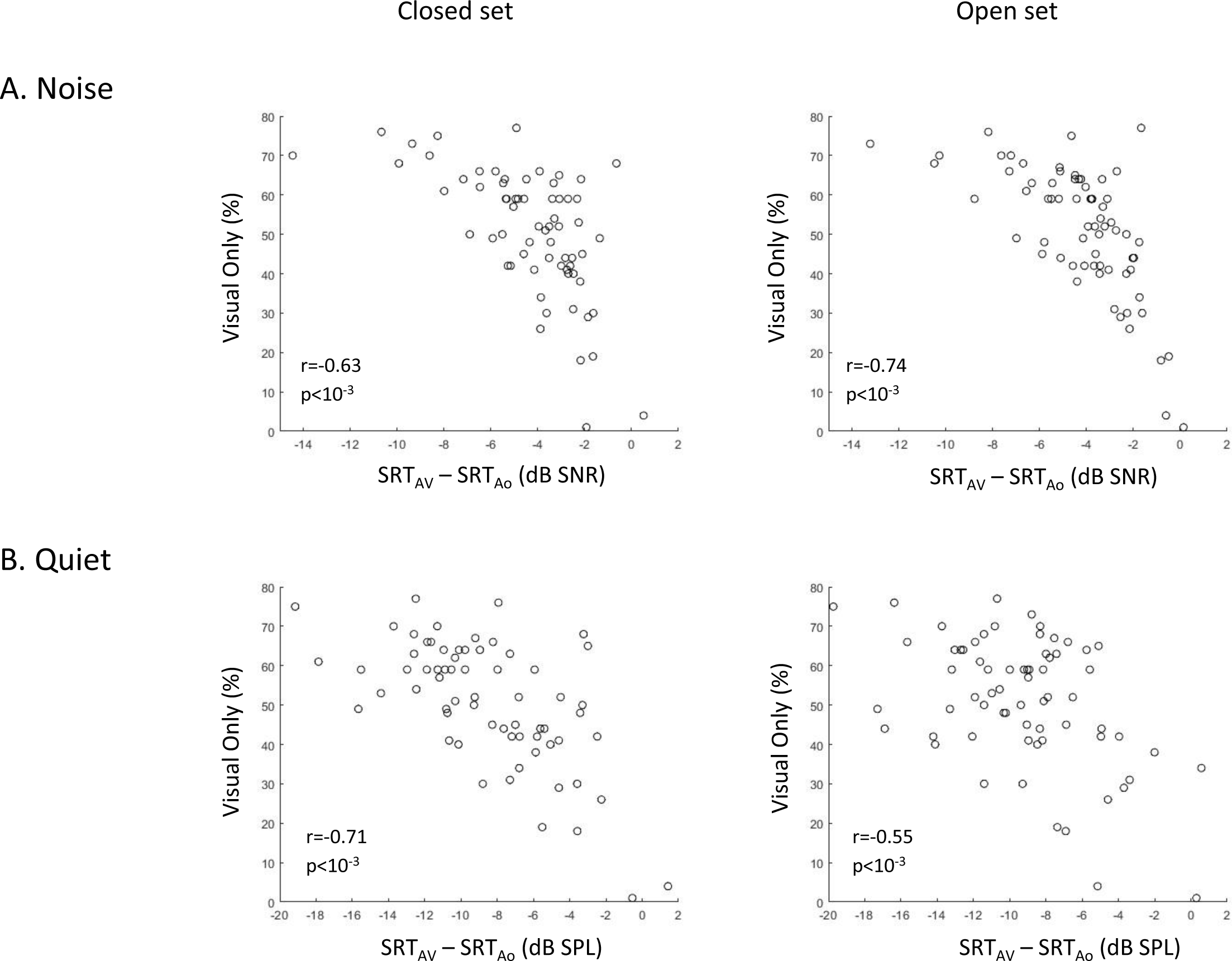
Visual Only scores as a function of the audio-visual benefit A. in noise (in dB SNR) and B. quiet (in dB SPL), in closed-set (left column) and open-set (right column) formats (individual data points) The scores of the test and retest sessions are dissociated, represented as two open circles per participant. r and p stand for the results of the Pearson correlations.

### Test-retest reliability

To assess the test-retest reliability of the audio-visual FrMST, we compared the within-participant standard deviations (test minus retest) to the between-participant standard deviations. In a homogenous group of young normal-hearing, the within- and between-participant standard deviations are expected to be similar in the Ao condition of the MST ^[31]^. In the AV and Vo conditions, a larger between-participant variability is expected ^[12]^. To evaluate the ability of the test to discriminate participants, we used the ‘2σ criterion’ ^[31]^. According to this criterion, the between-participant standard deviations should be double or more than the within-participant standard deviations to be able to discriminate between participants. ^[31]^

Figure 8 shows the within- and between-participant standard deviations of the scores in the conditions tested (SRTs and percentage of correct words for the Vo condition). The black line represents the 2σ criterion, which is the double of the within-participant standard deviation. Except for the Vo condition, none of the conditions exceeded the 2σ criterion, as found in Llorach et al. ^[12]^, who performed the same analysis. These results mean that it is possible to differentiate skilled from unskilled young normal-hearing participants in the Vo condition but not in the other conditions when using a 20-sentence list.

**Figure 8:**
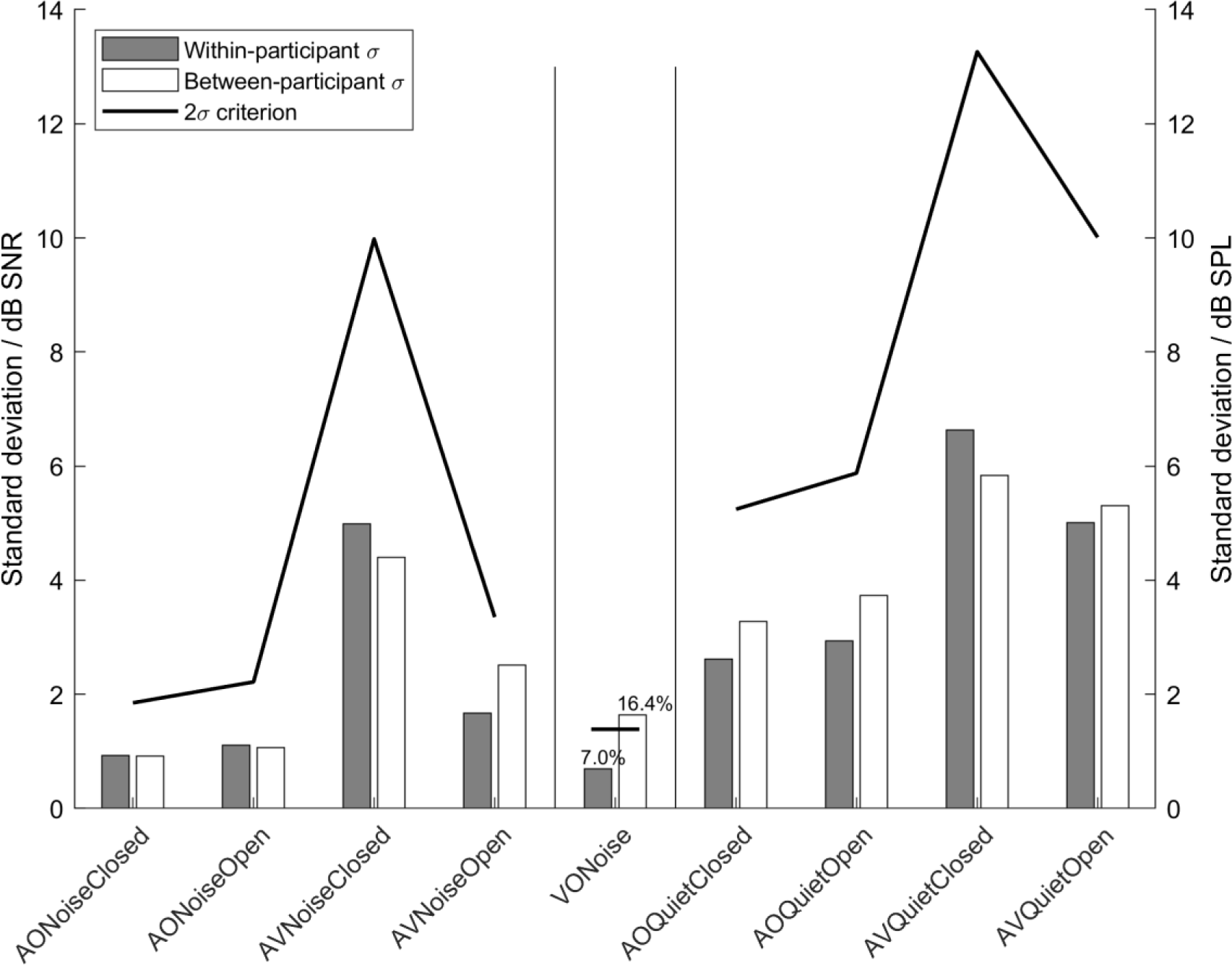
Within-participant (grey bars) and between-participant (white bars) standard deviations (SD) for all conditions. The 2σ criterion is indicated as a thick black line. On the left, SDs of speech in noise conditions expressed in dB SNR; middle, SDs of the visual-only, expressed in percentage; right, SDs of speech in quiet, expressed in dB SPL.

## Discussion

The present study designed and tested a standardized audio-visual material to evaluate lipreading skills and audio-visual benefit in a young normal-hearing sample. We used the French audio MST corpus validated by Jansen et al. ^[17]^, and the methodology described in Llorach et al. ^[12]^. We compare our results with these two studies.

### Population

The samples of the studies were similar in terms of age (mean 24.4 +/-2.8 years) and gender balance. We tested 8 more participants than Llorach et al. ^[12]^ and 5 more participants than Jansen et al. ^[17]^. They all had normal hearing.

### SRT50

A difference of −1.75 dB SNR was observed between the current study (−7.75 ± 0.79 dB SNR) and Jansen et al. ^[17]^ (−6 ± 0.6 dB SNR) when measuring SRT50 in audio-only and closed-set format. This outperformance may be explained by differences in calibration and technical configuration (binaural *versus* monaural presentation), and/or timing of the SRT50 assessment during the second session (greater training).

### Validation of the visual material

The absolute AV benefit (AV - Ao) was 4.6 dB SNR in noise and 9.25 dB SPL in quiet in the present study, *versus* 5.0 dB SNR and 7.0 dB SPL in Llorach et al. ^[12]^, and 3 dB SNR in Van de Rijt et al. ^[13]^. SRT differences in the AV and Ao conditions may show that the test was more difficult in French than in German. Similar differences between these languages have been mentioned before for SRT50 ^[32]^.

Although the AV benefit was in accordance with the literature and no participant reported AV lags, it is worth noting that our audio-visual material has inherent misalignments due to the recording method. The median and mean misalignment scores were however acceptable, around 32 ms . These pitfalls should be taken into account if audio-visual synchrony is paramount. Future work may use the warping path from the DTW to secondarily time-align the video recordings with the original sentences if more synchrony is required.

During the Visual-only condition, some participants scored up to 77% of comprehension. The average was 51.3% (±16.2%), similar to that of Llorach et al. ^[12]^ (50% ± 21.4%, range: 0%-84%).

### Training effect

An improvement of 1.8LdB SNR between the first and the sixth list is expected in the audio-only French MST, i.e., for SRT50 ^[17]^. Here, we found a 4.5 dB SNR improvement for the SRT80 in the AV condition in noise, between the first training list and the test list. This greater effect probably arose from the participants learning to lipread the material. Indeed, Lander & Davies ^[34]^ demonstrated that lipreading performance increased overall with practice. Accordingly, there was an average intelligibility improvement in the Vo only condition between the test and the retest session of 4.8% in the present study and 6.1% in Llorach et al. ^[12]^.

The training effect in the AV condition in noise statistically disappeared after the two first training lists. During the second training session, participants retained similar SRT scores as the one obtained for the fourth list of the first session, similarly to Llorach et al. ^[12]^.

### Test-retest reliability

We found a small within- and between-participants variability in the Audio-only in noise conditions, which was expected from a homogenous group of young normal-hearing participants. This variability increased in quiet conditions, in accordance with the literature_[12,35,36_].

The Visual-only scores were highly variable across participants, as expected ^[12,13,24]^. According to the ‘2σ criterion’, the Vo test is a reliable choice to determine if a participant is proficient in lipreading. In other words, it is enough to test a 20-sentence list in the Vo condition to determine the lipreading capabilities of an individual (after training with two AV lists).

Unfortunately, this was not the case for the AV conditions. The ‘2σ criterion’ was not fulfilled for any of the AV conditions, meaning that one 20-sentence list was not enough to differentiate AV speech perception between young NH participants and that several lists might be required to obtain a more precise score. These results are in accordance with Llorach et al. ^[12]^, where inherent asynchronies in the AV conditions were also present. These asynchronies could explain the increase in the variability in the AV measurements. Alternatively, it is plausible that audio-visual integration operates as an independent process^[37]^ exhibiting variability that is unrelated to the variability observed in the Ao and Vo conditions. To shed light on these hypotheses, future research should investigate the within-participant variability of audio-visual speech perception.

A larger between and within participants’ variability was found in the AV conditions in the present study as compared to Llorach et al. ^[12]^. This variability can be explained by the different individual lipreading abilities between the samples.

### Response format

There was no difference between the response formats once participants had been trained, similarly to the audio-visual German MST ^[12]^. For most languages, a closed-set response format means smaller SRTs (i.e., easier task), but not for German and Polish, as reported in Kollmeier et al. ^[32]^.

## Conclusion

We have created an audio-visual version of the French version of the MST by visually dubbing the original validated audio material.

The SRT80 values for young normal hearing participants in the AV condition were −9.2LdB SNR in noise and 16.8LdB SPL in quiet. The absolute AV benefit was 4.6 dB SNR in noise and 9.25 dB SPL in quiet. Lipreading scores (visual-only sentences) ranged from 1% to 77%. These results are in accordance with those already published for AV MSTs in other languages.

One should take into account that the audio-visual test suffers from ceiling effects as some participants were able to recognize more than 50% of the words in the Visual-only situation. Therefore, targeting SRT80 is recommended. Due to the within-participant variability of the AV test more than one list might be required to obtain a precise auditory-visual score when comparing young normal-hearing participants.

This audio-visual material can be used for clinical purposes, but in practice, it is not possible to carry out all the conditions described, especially in hearing-impaired participants. For clinical purposes, we suggest performing two AV training lists in noise (closed-set or open-set format, not significant difference in French), followed by: one list in audio-visual condition in noise, one list in audio only condition in noise, and one list in visual only condition in noise, in a randomised order.

Further research will evaluate the Audio-visual FrMST in hearing-impaired cohorts, and especially their audio-visual gain in quiet and in noise.

## Data Availability

All data produced in the present study are available upon reasonable request to the authors

## Acknowledgements

We thank Marie-Amélie Bizouard and Gregory Gerenton for their technical help, Jan Wouters and Volker Hohmann for relevant discussions. This work was funded by Fondation Pour l’Audition FPA RD-2020-10 (for LLR, TD, LHA, DL) and by the Deutsche Forschungsgemeinschaft (for GL, DFG, Cluster of Excellence EXC 1077/1 “Hearing4all”, and SFB1330 Projects B1 and C4). We thank one of the reviewers for relevant discussions about the DTW calculations, which helped improve our work.

Statements: The video recordings of the female French Matrix Sentence Test can be found free of charge online (https://doi.org/10.5281/zenodo.8188917). Please cite the present article and the following when using the material:

Jansen, S., Luts, H., Wagener, K. C., Kollmeier, B., Del Rio, M., Dauman, R., James, C., Fraysse, B., Vormès, E., Frachet, B., Wouters, J., & van Wieringen, A. (2012). Comparison of three types of French speech-in-noise tests: A multi-center study. *International Journal of Audiology*, *51*(3), Art. 3. https://doi.org/10.3109/14992027.2011.633568

Llorach, G., Kirschner, F., Grimm, G., Zokoll, M. A., Wagener, K. C., & Hohmann, V. (2022). Development and evaluation of video recordings for the OLSA matrix sentence test. *International Journal of Audiology*, *61*(4), 311L321. https://doi.org/10.1080/14992027.2021.1930205

The audio recordings are not provided in the dataset, but research licenses are available from Hörzentrum Oldenburg gGmbH. Please refer to Hörzentrum Oldenburg gGmbH for the audio material (https://www.hz-ol.de/en/matrix.html).

The software that links audio and video in the design process is available open-source at GitHub - gerardllorach/audiovisualdubbedMST. This is a repository with guidelines and code to create visual material for Matrix Sentence Test for speech audiometry. https://github.com/gerardllorach/audiovisualdubbedMST

The research software that runs the test is available for collaborative work with Hörzentrum Oldenburg gGmbH. Please contact.

## References

1. Dell’Aringa, A., Adachi, E., & Dell’Aringa, A. (2007). Lip reading role in the hearing aid fitting process. Brazilian journal of otorhinolaryngology, 73, 95L99. 10.1016/S1808-8694(15)31129-0

2. Pimperton, H., Ralph-Lewis, A., & MacSweeney, M. (2017). Speechreading in Deaf Adults with Cochlear ImplantsL: Evidence for Perceptual Compensation. Frontiers in Psychology, 8. https://www.frontiersin.org/article/10.3389/fpsyg.2017.00106

3. Lazard, D. S., & Giraud, A.-L. (2017). Faster phonological processing and right occipito-temporal coupling in deaf adults signal poor cochlear implant outcome. Nature Communications, 8(1), Article 1. 10.1038/ncomms14872

4. Bottalico, P., Murgia, S., Puglisi, G. E., Astolfi, A., & Kirk, K. I. (2020). Effect of masks on speech intelligibility in auralized classrooms. The Journal of the Acoustical Society of America, 148(5), Article 5. 10.1121/10.0002450

5. Sönnichsen, R., Llorach Tó, G., Hochmuth, S., Hohmann, V., & Radeloff, A. (2022). How Face Masks Interfere With Speech Understanding of Normal-Hearing IndividualsL: Vision Makes the Difference. Otology & Neurotology: Official Publication of the American Otological Society, American Neurotology Society [and] European Academy of Otology and Neurotology, 43(3), Article 3. 10.1097/MAO.0000000000003458

6. Tofanelli, M., Capriotti, V., Gatto, A., Boscolo-Rizzo, P., Rizzo, S., & Tirelli, G. (2022). COVID-19 and DeafnessL: Impact of Face Masks on Speech Perception. Journal of the American Academy of Audiology, 33(2), 98L104. 10.1055/s-0041-1736577

7. Yi, H., Pingsterhaus, A., & Song, W. (2021). Effects of Wearing Face Masks While Using Different Speaking Styles in Noise on Speech Intelligibility During the COVID-19 Pandemic. Frontiers in Psychology, 12. https://www.frontiersin.org/article/10.3389/fpsyg.2021.682677

8. Arnal, L. H., Morillon, B., Kell, C. A., & Giraud, A.-L. (2009). Dual neural routing of visual facilitation in speech processing. Journal of Neuroscience, 29(43), Article 43.

9. Bourguignon, M., Baart, M., Kapnoula, E. C., & Molinaro, N. (2020). Lip-reading enables the brain to synthesize auditory features of unknown silent speech. The Journal of Neuroscience, 40(5), 1053L1065. 10.1523/JNEUROSCI.1101-19.2019

10. Erber, N. P. (1975). Auditory-visual perception of speech. The Journal of Speech and Hearing Disorders, 40(4), 481L492. 10.1044/jshd.4004.481

11. Kessous, L., Castellano, G., & Caridakis, G. (2010). Multimodal emotion recognition in speech-based interaction using facial expression, body gesture and acoustic analysis. Journal on Multimodal User Interfaces, 3(1L2), 33L48. 10.1007/s12193-009-0025-5

12. Llorach, G., Kirschner, F., Grimm, G., Zokoll, M. A., Wagener, K. C., & Hohmann, V. (2022). Development and evaluation of video recordings for the OLSA matrix sentence test. International Journal of Audiology, 61(4), 311L321. 10.1080/14992027.2021.1930205

13. van de Rijt, L. P. H., Roye, A., Mylanus, E. A. M., van Opstal, A. J., & van Wanrooij, M. M. (2019). The Principle of Inverse Effectiveness in Audiovisual Speech Perception. Frontiers in Human Neuroscience, 13, 335. 10.3389/fnhum.2019.00335

14. Fournier, J.-E., & Aubin, A. P. (1951). Audiométrie vocalelJ: Les épreuves d’intelligibilité et leurs applications au diagnostic, à l’expertise et à la correction prothétique des surdités. Maloine.

15. Lafon, J. C. (1972). Phonetic test, phonation, audition. JFORL. Journal francais d"oto-rhino-laryngologie; audiophonologie et chirurgie maxillo-faciale, 21(3), 223L229. Scopus.

16. Zimpfer, V., Andéol, G., Blanck, G., Suied, C., & Fux, T. (2020). Development of a French version of the Modified Rhyme Test. The Journal of the Acoustical Society of America, 147(1), EL55LEL61. 10.1121/10.0000559

17. Jansen, S., Luts, H., Wagener, K. C., Kollmeier, B., Del Rio, M., Dauman, R., James, C., Fraysse, B., Vormès, E., Frachet, B., Wouters, J., & van Wieringen, A. (2012). Comparison of three types of French speech-in-noise testsL: A multi-center study. International Journal of Audiology, 51(3), Article 3. 10.3109/14992027.2011.633568

18. Leclercq, F., Renard, C., & Vincent, C. (2018). Speech audiometry in noiseL: Development of the French-language VRB (vocale rapide dans le bruit) test. European Annals of Otorhinolaryngology, Head and Neck Diseases, 135(5), Article 5. 10.1016/j.anorl.2018.07.002

19. Luck, S. J. (2014). An Introduction to the Event-Related Potential Technique, second edition. MIT Press.

20. Brand, T., Kissner, S., Jürgens, T., Berg, D., & Kollmeier, B. (2011, mars 5). Adaptive Algorithmen zur Bestimmung der 80%-Sprachverständlichkeitsschwelle.

21. Brand, T., & Kollmeier, B. (2002). Efficient adaptive procedures for threshold and concurrent slope estimates for psychophysics and speech intelligibility tests. The Journal of the Acoustical Society of America, 111(6), Article 6. 10.1121/1.1479152

22. A. O’Beirne, G. A., Trounson, R. H., McClelland, A. D., Jamaluddin, S. A., & Maclagan, M. (2015). Development of an auditory-visual matrix sentence test in New Zealand English. Journal of International Advanced Otology, 11(Supplement 1), Article Supplement 1.

22. Trounson, R. H. (2012). Development of the UC Auditory-Visual Matrix Sentence Test.

23. Jamaluddin, S. A. (2016). Development and evaluation of the digit triplet and auditory-visual matrix sentence tests in Malay.

24. Hochmuth, S., Kollmeier, B., Brand, T., & Jürgens, T. (2015). Influence of noise type on speech reception thresholds across four languages measured with matrix sentence tests. International Journal of Audiology, 54(sup2), Article sup2. 10.3109/14992027.2015.1046502

25. Moore, B. C. J. (2012). An Introduction to the Psychology of Hearing. BRILL.

26. Schubotz, W., Brand, T., Kollmeier, B., & Ewert, S. D. (2016). Monaural speech intelligibility and detection in maskers with varying amounts of spectro-temporal speech features. The Journal of the Acoustical Society of America, 140(1), Article 1. 10.1121/1.4955079

27. Bernstein, L., Auer, E., & Takayanagi, S. (2004). Auditory speech detection in noise enhanced by lipreading. Speech Communication, 44, 5L18. 10.1016/j.specom.2004.10.011

28. Ross, L. A., Saint-Amour, D., Leavitt, V. M., Javitt, D. C., & Foxe, J. J. (2007). Do you see what I am saying? Exploring visual enhancement of speech comprehension in noisy environments. Cerebral Cortex (New York, N.Y.: 1991), 17(5), 1147L1153. 10.1093/cercor/bhl024

29. Sumby, W. H., & Pollack, I. (2005). Visual Contribution to Speech Intelligibility in Noise. The Journal of the Acoustical Society of America, 26(2), 212L215. 10.1121/1.1907309

30. Wagener, K. C., & Brand, T. (2005). Sentence intelligibility in noise for listeners with normal hearing and hearing impairmentL: Influence of measurement procedure and masking parameters. International Journal of Audiology, 44(3), Article 3. 10.1080/14992020500057517

31. Kollmeier, B., Warzybok, A., Hochmuth, S., Zokoll, M. A., Uslar, V., Brand, T., & Wagener, K. C. (2015). The multilingual matrix testL: Principles, applications, and comparison across languages: A review. International Journal of Audiology, 54 Suppl 2, 3L16. 10.3109/14992027.2015.1020971

32. Yakel, D. A., Rosenblum, L. D., & Fortier, M. A. (2000). Effects of talker variability on speechreading. Perception & Psychophysics, 62(7), Article 7. 10.3758/BF03212142

33. Lander, K., & Davies, R. (2008). Does face familiarity influence speechreadability? Quarterly Journal of Experimental Psychology, 61(7), Article 7. 10.1080/17470210801908476

34. Smoorenburg, G. F. (1992). Speech reception in quiet and in noisy conditions by individuals with noiseLinduced hearing loss in relation to their tone audiogram. The Journal of the Acoustical Society of America, 91(1), 421L437. 10.1121/1.402729

35. Souza, P. E., Boike, K. T., Witherell, K., & Tremblay, K. (2007). Prediction of Speech Recognition from Audibility in Older Listeners with Hearing LossL: Effects of Age, Amplification, and Background Noise. Journal of the American Academy of Audiology, 18(01), 054L065. 10.3766/jaaa.18.1.5

36. Grant, K. W. (2002). Measures of auditory-visual integration for speech understandingL: A theoretical perspective. Journal of the Acoustical Society of America, 112(1), 30L33. 10.1121/1.1482076

37. Sakoe, H., & Chiba, S. (1978). Dynamic programming algorithm optimization for spoken word recognition. IEEE Transactions on Acoustics, Speech, and Signal Processing, 26(1), Article 1. 10.1109/TASSP.1978.1163055

